# Single cell analysis of menstrual endometrial tissues defines phenotypes associated with endometriosis

**DOI:** 10.1101/2022.02.10.22270810

**Authors:** Andrew J. Shih, Robert P. Adelson, Himanshu Vashistha, Houman Khalili, Ashima Nayyar, Radha Puran, Rixsi Herrera, Prodyot K. Chatterjee, Annette T. Lee, Alexander M. Truskinovsky, Kristine Elmaliki, Margaret DeFranco, Christine N. Metz, Peter K. Gregersen

## Abstract

**Background:** Endometriosis is a common, complex disorder which is underrecognized and subject to prolonged delays in diagnosis. It is accompanied by significant changes in the eutopic endometrial lining.

**Methods:** We have undertaken the first single cell RNA-sequencing (scRNA-Seq) comparison of endometrial tissues in freshly collected menstrual effluent (ME) from 33 subjects, including confirmed endometriosis patients (cases) and controls as well as symptomatic subjects (who have chronic symptoms suggestive of endometriosis but have not been diagnosed).

Results

We identify a unique subcluster of proliferating uterine natural killer (uNK) cells in ME-tissues from controls that is almost absent from endometriosis cases, along with a striking reduction of total uNK cells in the ME of cases (p<10^-16^). In addition, an IGFBP1+ decidualized subset of endometrial stromal cells are abundant in the shed endometrium of controls when compared to cases (p<10^-16^) confirming findings of compromised decidualization of cultured stromal cells from cases. By contrast, endometrial stromal cells from cases are enriched in cells expressing pro-inflammatory and senescent phenotypes. An enrichment of B cells in the cases (p=5.8 × 10^-6^) raises the possibility that some may have chronic endometritis, a disorder which predisposes to endometriosis.

**Conclusions:** We propose that characterization of endometrial tissues in ME will provide an effective screening tool for identifying endometriosis in patients with chronic symptoms suggestive of this disorder. This constitutes a major advance, since delayed diagnosis for many years is a major clinical problem in the evaluation of these patients. Comprehensive analysis of ME is expected to lead to new diagnostic and therapeutic approaches to endometriosis and other associated reproductive disorders such as female infertility.

## Background

Endometriosis is a common and heterogeneous disorder that is characterized by the growth of endometrial-like tissues outside of the uterus, most commonly in the peritoneal cavity and associated with inflammation (1). While the pathogenesis of endometriosis is not understood, retrograde menstruation of endometrial cells and tissues via the fallopian tubes is one accepted theory for the development of endometriosis lesions in the peritoneal cavity (2, 3). However, retrograde menstruation occurs in nearly all women (4), yet endometriosis occurs in approximately one in ten females in their reproductive years (3). Thus, other factors must contribute to the development of endometriosis. While there is a significant genetic component to endometriosis (5), very little is known about how these putative risk alleles function. On the other hand, the eutopic endometrium of patients with endometriosis is significantly different when compared to the endometrium of those without endometriosis, with inflammatory changes noted in the setting of endometriosis(6-9). We have undertaken a detailed analysis of endometrial tissues and cells present in menstrual effluent (ME), since ME is the critical biological sample transferred to the pelvic cavity, where most endometriosis lesions grow.

Most previous investigations of ME have involved the phenotypic analysis by immunofluorescence, flow cytometry, and/or *in vitro* culture of single cell suspensions collected using menstrual cups (10-14). Our previous flow cytometry studies showed that uterine natural killer (uNK) cells were relatively depleted in ME from endometriosis cases vs. controls (11).

However, this study was limited by the analysis of relatively few cell types in ME, with no assessment of specific subsets of cells. In addition, we demonstrated a defect in decidualization capacity of endometrial stromal cells grown from the ME of patients with endometriosis when compared to ME-stromal cells grown from healthy controls (11, 15). While these earlier results potentially provided a basis for a screening test for endometriosis, these analyses relied on laborious and expensive cell culture and *in vitro* assays, making them impractical for clinical application.

Herein we investigated fresh ME as an unexplored and important biological specimen for the development of non-invasive diagnostics based on the direct analysis of endometrial tissue fragments. We show that ME contains large numbers of shed fragments from endometrial tissues. Using enzymatic digestion of ME and associated tissues followed by single cell RNA-sequencing (scRNA-Seq) analysis, we compared the major cellular differences and gene expression profiles found in ME collected from healthy controls (without symptoms of endometriosis) and patients diagnosed with endometriosis (confirmed by laparoscopic surgery with positive confirmation by pathology), as well as patients with symptoms of endometriosis (e.g., recurrent dysmenorrhea, persistent abdominal bloating, dyspareunia, dysuria, and/or dyschezia) who are not yet diagnosed. In order to gain insight into the pathogenesis of endometriosis, we particularly focused on the phenotypes of stromal and uNK cells in ME through scRNA-Seq because these are abundant and have been previously shown to be abnormal in eutopic endometrium of patients.

## Methods

### Human subjects and menstrual effluent collections

Menstrual effluent (ME) was collected as previously described (11, 15). Briefly, women of reproductive age (N=33, age 20-45 years, average age 33.6 years) living in North America who were not pregnant or breastfeeding, who were menstruating, and who were willing to provide ME samples were recruited mainly via social media and consented to the ROSE study (IRB#13-376A) (https://feinstein.northwell.edu/institutes-researchers/institute-molecular-medicine/robert-s-boas-center-for-genomics-and-human-genetics/rose-research-outsmarts-endometriosis).

Women with histologically confirmed endometriosis (determined following excision laparoscopic surgery and documented in a pathology report without revised American Society for Reproductive Medicine (rASRM) staging/classification) were enrolled as ‘endometriosis’ subjects (N=11). Women who reported chronic symptoms consistent with endometriosis (e.g., recurrent dysmenorrhea, persistent abdominal bloating, dyspareunia, dysuria, and/or dyschezia), but not yet diagnosed with endometriosis (or not) were enrolled as ‘symptomatic’ subjects (N=13). Control subjects living in North America who self-reported no gynecologic history suggestive of a diagnosis of endometriosis (and the absence of polycystic ovarian syndrome, and pelvic inflammatory disease) were recruited mainly via social media and enrolled as ‘controls’ (N=9).

Endometriosis, symptomatic, and control subjects collected their ME using an ‘at home’ ME collection kit for 4-8 hours on the day of their heaviest menstrual flow (typically day 1 or 2 of the cycle) with a menstrual cup (provided by DIVA International), except for one subject who collected ME using a novel menstrual collection sponge (as previously described (15)). After collection, ME was shipped priority overnight at °C to the laboratory for processing. ME collected from menstrual cups was mixed 1:1 with DMEM for processing. For the saturated menstrual collection sponge, ME tissue was collected after rinsing the sponges with PBS to collect cells and tissue. Demographic and gynecologic/health data (including hormone usage, menstrual cycle information, and pain/pain medications) for controls, endometriosis subjects, and symptomatic subjects (and the total cohort) are shown in Table 1.

**Table 1.** Subject Group Characteristics – Control (CTRL), Dx (Diagnosed), Sx (Symptomatic

|  | **CTRL** | **Dx** | **Sx** | **TOTAL** | **P value (CTRL vs Dx)** |
| --- | --- | --- | --- | --- | --- |
| **Age (years)**<br>(mean±SD) | 33.4±5.4 | 35.2±4.4 | 32.3±8.2 | 33.6±6.3 | 0.42 |
| **BMI (kg/m<sup>2</sup>)**<br>(mean±SD) | 24.2±7.1 | 28.4±5.4 | 26.8±7.6 | 26.5±6.8 | 0.15 |
| **Age at menarche (years)**<br>(mean±SD) | 12.0±0.9 | 11.6±1.6 | 12.6±1.7 | 12.0±1.5 | 0.51 |
| **Race/ethnicity** |  |  |  |  | 0.34 |
| Caucasian | 7/9 (78%) | 0/11 (91%) | 13/13 (100%) | 30/33 (91%) |  |
| Black | 1/9 (11%) | 0/11 (0%) | 0/13 (0%) | 1/33 (3%) |  |
| Mixed | 1/9 (11%) | 0/11 (0%) | 0/13 (0%) | 1/33 (3%) |  |
| Other | 0/9 (0%) | 1/11 (9%) | 0/13 (0%) | 1/33 (3%) |  |
| Hispanic | 0/9 (0%) | 0/11 (0%) | 1/13 (8%) | 1/33 (3%) |  |
| **Typical cycle length (days)** |  |  |  |  | 0.52 |
| 21-25 days | 0/9 (0%) | 1/11 (9%) | 3/13 (23%) | 4/33 (12%) |  |
| 26-31 days | 8/9 (89%) | 7/11 (64%) | 8/13 (62%) | 23/33 (70%) |  |
| 32-39 days | 1/9 (11%) | 2/11 (18%) | 2/13 (15%) | 5/33 (15%) |  |
| >40 days | 0/9 (0%) | 1/11 (9%) | 0/13 (0%) | 1/33 (3%) |  |
| **Typical bleed time (days)** |  |  |  |  | 0.36 |
| <3 d | 0/9 (0%) | 2/11 (18%) | 0/13 (0%) | 2/33 (6%) |  |
| 3-5 d | 5/9 (56%) | 6/11 (55%) | 8/13 (62%) | 19/33 (58%) |  |
| 6-8 d | 4/9 (44%) | 3/11 (23%) | 5/13 (38%) | 12/33 (36%) |  |
| **Typical flow** |  |  |  |  | 0.96 |
| Light | 1/9 (11%) | 1/11 (9%) | 0/13 (0%) | 2/33 (6%) |  |
| Moderate | 2/9 (22%) | 2/11 (18%) | 3/13 (23%) | 7/33 (21%) |  |
| Moderately Heavy | 3/9 (33%) | 5/11 (45%) | 9/13 (69%) | 17/33 (52%) |  |
| Heavy | 3/9 (33%) | 3/11 (23%) | 1/13 (8%) | 7/33 (21%) |  |
| **Hormone use** |  |  |  |  | 0.35 |
| Yes | 0/9 (0%) | 1/11 (9%) | 1/13 (8%) | 2/33 (6%) |  |
| **Pain in this cycle** |  |  |  |  | 0.06 |
| Yes | 5/9 (56%) | 10/11 (91%) | 13/13 (100%)* | 28/33 (85%) |  |
| None | 4/9 (44%) | 1/11 (9%) | 0/13 (0%) | 5/33 (15%) |  |
| Mild | 2/9 (22%) | 3/11 (23%) | 3/13 (23%) | 8/33 (24%) |  |
| Moderate | 3/9 (33%) | 6/11 (55%) | 4/13 (31%) | 13/33 (40%) |  |
| Severe | 0/9 (0%) | 1/11 (9%) | 6/13 (46%) | 7/33 (21%) |  |
| **Pain medication in this cycle**<br>(Midol, Advil, Tylenol, Naproxen, Hydromorphone 2mg/Baclofen/diazepam/Ketamine 8/10/15mg) |  |  |  |  |  |
| Yes | 1/9 (11%) | 6/11 (55%) | 10/13 (77%) | 17/33 (52%) | 0.04 |

No comparisons of CTRL vs. Sx were significant for the above characteristics, except pain (yes/no):
\*P=0.008

### Immunostaining of ME-derived tissue fragments

ME-derived tissue fragments were obtained from controls, symptomatic subjects, and endometriosis patients (n=2 each); tissue fragments were collected by pouring ME over a 70µ filter, fixed, and transferred to the clinical pathology lab for paraffin embedding and hematoxylin and eosin (H&E) staining. CD10 was chosen for immunohistochemical analysis because it is a sensitive marker of eutopic endometrial stroma (16) and because adjunctive use of CD10 immunostaining with H&E staining enhances the histologic detection of endometriosis (17). CD56 was chosen because uNK cells stain brightly with CD56. H&E slides and immunostained slides were examined microscopically and imaged by a pathologist. Representative images are shown in Fig. 1.

**Fig. 1.**
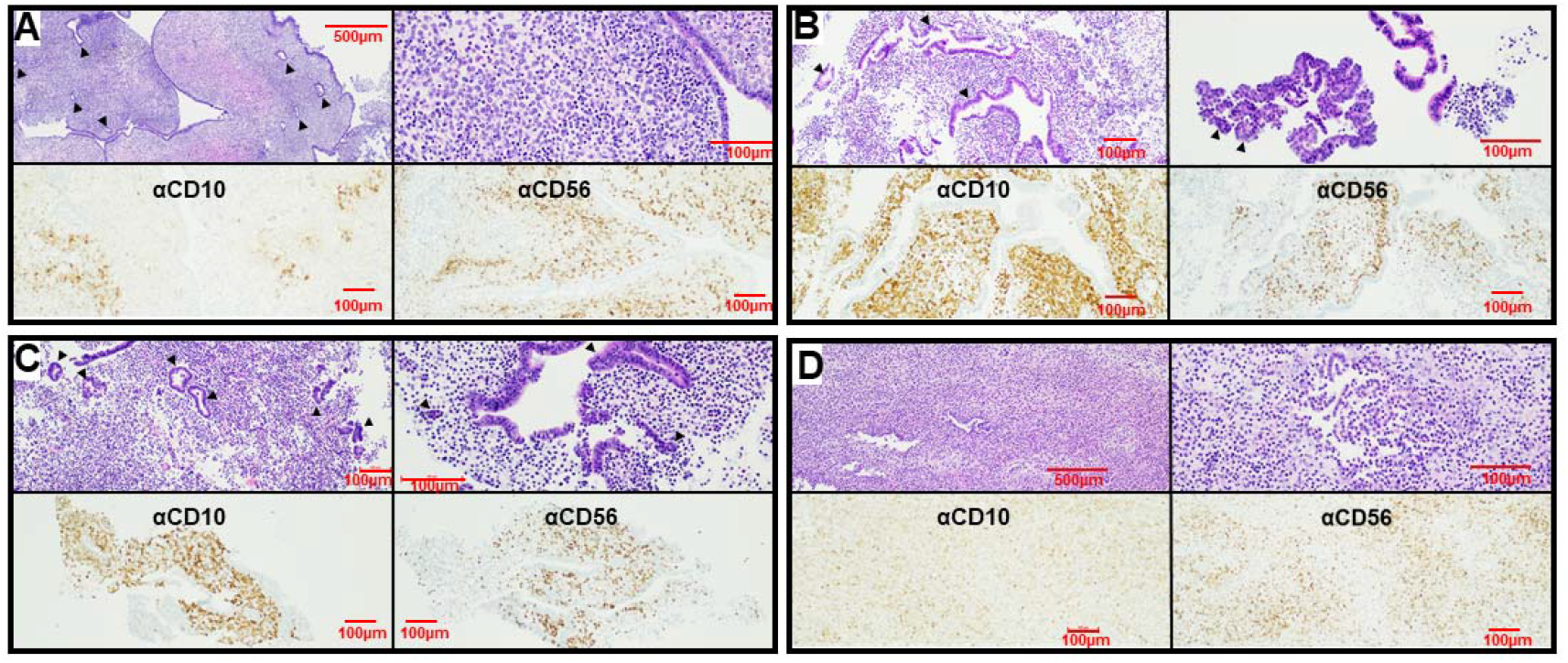
*ME contains endometrial tissues.* Histological analysis of endometrial tissues isolated from the menstrual effluent (ME) from 4 separate subjects: A) control subject, B-C) two subjects with pathologically confirmed endometriosis, and D) subject chronic symptoms of endometriosis (not yet diagnosed). Upper panels for A-D: H&E staining is shown in two panels at two magnifications for each individual: A) 40X (left) and 200X (right); B) 100X and 200X; C) 100X and 200X; and D) 40X and 200X; arrowheads point to glandular epithelium. Sections show typical late secretory/menstrual endometrium with expanded stroma containing scattered inflammatory cells and secretory and inactive type glands. Lower panels for A-D: immunostaining with anti-CD10 and anti-CD56 antibodies to detect stromal cells (left) and uterine NK (uNK) cells (right), respectively, at 100X. Scale bars are shown in each image.

### Processing menstrual effluent for scRNA-Seq analyses

Whole (unfractionated) ME (2.5-10ml) was digested with Collagenase I (1mg/ml, Worthington Biochemical Corporation, Lakewood, NJ) and DNase I (0.25mg/ml, Worthington Biochemical Corporation) at 37°C for 10-30 min using the gentleMACS™ Tissue Octo Dissociator (Miltenyi Biotec, Cambridge, MA) using C tubes and Program 37CMulti_E_01 (31 min). After digestion, the sample was sieved over a 70µ filter and washed with DMEM 10% fetal bovine serum (FBS) to neutralize digestion enzymes; the flow through was sieved over a 40µ filter and washed with DMEM 10%FBS. After collecting the single cells (from the flow through) following centrifugation (350xg for 5 min), Neutrophils were removed using the EasySep™ HLA Chimerism Whole Blood CD66b Positive Selection Kit (STEMCELL, Cambridge, MA), according to the manufacturer’s protocol. The neutrophil pellet was frozen at -80°C and used as a source of subject DNA for genotyping (see below). The resultant cells were depleted of red blood cells using the EasySep™ RBC Depletion Reagent (STEMCELL), according to the manufacturer’s protocol, and then washed and subjected to density gradient centrifugation using Ficoll-Paque PLUS (Sigma-Aldrich, St. Louis, MO) to collect mononuclear cells, according to manufacturer’s directions. To collect ME-tissue, whole ME (2.5-10ml) was sieved over a 70µ filter and washed with DMEM; the ME-tissues trapped on the filter was collected and digested with Collagenase I (1mg/ml, Worthington Biochemical Corporation, Lakewood, NJ) and DNase I (0.25mg/ml, Worthington Biochemical Corporation) at 37°C for 10 min and processed as described above for whole ME, except without a density gradient centrifugation step. The resultant whole ME cells were enumerated, and viability was assessed using ViaStain™ AOPI Staining Solution and the Nexcelom Cellometer (Lawrence, MA). Preparations with >80% viability were processed for scRNA-Seq. Cells were immediately fixed in methanol for scRNA-Seq, as described by Chen for peripheral blood mononuclear cells (18)). Briefly, cells were washed and resuspended in a 200µl Ca^++^ and Mg^++^-free PBS, followed by dropwise addition of chilled 100% methanol (800µl, final 80% w/v). Fixed cells were stored at -20°C for 20min and then stored at -80°C until used for scRNA-Seq. A pilot experiment was performed with a single ME sample, which was processed and either prepared immediately for scRNA-Seq (without methanol fixation and freezing) or was fixed in methanol and frozen, as described above to optimize our scRNA-Seq methods. The data showed almost identical scRNA-Seq results using both methods (see Additional File 1: Fig. S1). Thus, all ME samples were methanol fixed and frozen, allowing for cost-effective ‘batching’ in scRNA-SEQ.

### Processing of samples for single cell sequencing

Methanol-fixed cells were removed from -80°C and placed on ice for 5 min before centrifugation (1000xg for 5 min). Methanol-PBS supernatant was completely removed and cells were rehydrated in 0.04% bovine serum albumin (BSA) + 1mM dithiothreitol (DTT) + 0.2 U/ul RNase Inhibitor in 3X SSC (saline sodium citrate buffer solution) Buffer (Sigma). An aliquot of fixed cells was stained with Trypan Blue and visualized under the microscope. The cells were counted and pooled from different donors at equal ratios, filtered using 35µ strainer (Falcon), recounted and brought up to a final conc. of 2,000 cells/µl and proceeded immediately for GEM generation and barcoding on a 10X Chromium using Next GEM 3’ v3.1 reagents (10X Genomics). Libraries were constructed following 10X Genomics’ recommendations and quality was assessed on a High Sensitivity DNA chip on a BioAnalyzer 2100 (Agilent) before loading (1.8 pM) and sequencing on an Illumina Nextseq 500 using a High Output kit v2.5 (150 cycles).

Five subjects were pooled together into a single 10x lane with at least one of each phenotype per run with a total of 8 runs for ME-tissue and 3 runs of whole ME. The ME-tissue runs had 44,135 total cells, of which 5,147 had ambiguous calls in Demuxlet, 2,632 were doublets and 36,356 were singlets; only the singlets were analyzed. The whole-ME runs had 30,090 total cells, of which 5,556 had ambiguous calls, 2,776 were doublets, and 21,758 singlets (and hence analyzed). A total of 43,054 cells were analyzed in this study following filtering and QC (thresholds of > 10% mitochondrial reads < 500 nUMI (number of unique molecular identifiers) or > 50000 nUMI or > 6000 unique features per cell).

### Single cell RNA-Sequencing and analyses and statistics

Samples were converted from raw bcl files to gene by cell matrices using CellRanger 6.0 aligned to 10x Genomics’ GRCh38-3.0.0 reference. Individuals were demultiplexed via Demuxlet (19) using genotypes taken from SNPs on the Illumina GSAv3 genotyping array, run on DNA prepared from neutrophils isolated from ME. The thresholds in Demuxlet were adjusted to the expected doublet rate and those marked as doublets were removed. Downstream analysis and visualization were done using Seurat 4.0 (20). Briefly, there were at least 25,000 reads per cell on average per 10x run and the mean number of genes captured was 1,388 (± 896 (mean ± standard deviation [SD]). There was no significant difference between the various clinical groups (controls, cases, symptomatic) in these values. Genes were filtered out if they were expressed in less than 3 cells while cells were filtered out if they had > 10% mitochondrial reads, 500 < nUMI < 50000 and > 6000 unique features. For the analysis of ME-tissue samples, only subjects with information on at least 500 cells per subject were retained. After filtering the cell yields were comparable in each group (mean ± SD: 1,256 ±732 and 1,319 ±767 in ME-tissue and whole ME, respectively). Gene expression normalization and cell clustering was done using the SCTransform pipeline (21) with percent mitochondrial reads regressed out and person specific batch effects corrected using Harmony (22). Identification of cell clusters was done using known marker genes (Additional File 2: Table S1) (23-30) with differential gene expression calculated using a Wilcoxon rank sum test. Enrichment of cell clusters of specific phenotypes was done using MASC (mixed-effects modeling of associations of single cells) (https://github.com/immunogenomics/masc), which essentially uses a percentage of cells per cluster while also taking into account technical covariates; 10x library batch, preparation (whole ME or ME-tissue), nUMI per cell, percent mitochondrial reads and phase are accounted for. All datasets are deposited in the National Center for Biotechnology Information/Gene Expression Omnibus (GEO) accession number GSE203191.

## Results

### Endometrial tissue fragments are present in fresh menstrual effluent

We carried out histological assessment of fresh menstrual effluent (ME)-associated tissues isolated from ME. Representative H&E sections of ME-derived tissue fragments from four subjects (1 control, 2 laparoscopically/histologically confirmed endometriosis subjects, and 1 symptomatic subject) show the presence of endometrial tissues with mucosal and glandular epithelium and areas of stroma. The endometrium had typical late secretory/menstrual morphology with expanded stroma containing scattered inflammatory cells, and secretory and inactive-type glands (Fig. 1A-D, upper panels). Immunostaining of ME-derived tissue sections reveals a range of stromal cells stained with antibodies to CD10, a clinically used marker of endometrial stroma (16, 17), and an abundance of uNK cells (stained with antibodies to CD56 (NCAM), an archetypical marker of NK cells [Fig. 1A-D, lower panels]).

### Single cell RNA sequencing (scRNA-Seq) of digested freshly processed ME reveals the presence of a heterogenous mixture of immune and non-immune cells

We have analyzed ME samples from 33 subjects, including age-matched healthy controls (N=9), endometriosis cases (N=11), and subjects with chronic symptoms suggestive of endometriosis but not yet diagnosed (N=13) (see Table 1). ME samples from either whole ME (unfractionated) or ME samples enriched for tissues (“ME-tissue”) were digested with collagenase I and DNase I, depleted of neutrophils, and processed for scRNA-Seq, as described in the methods. As shown in Fig. 2 a graph-based clustering approach using

**Fig. 2.**
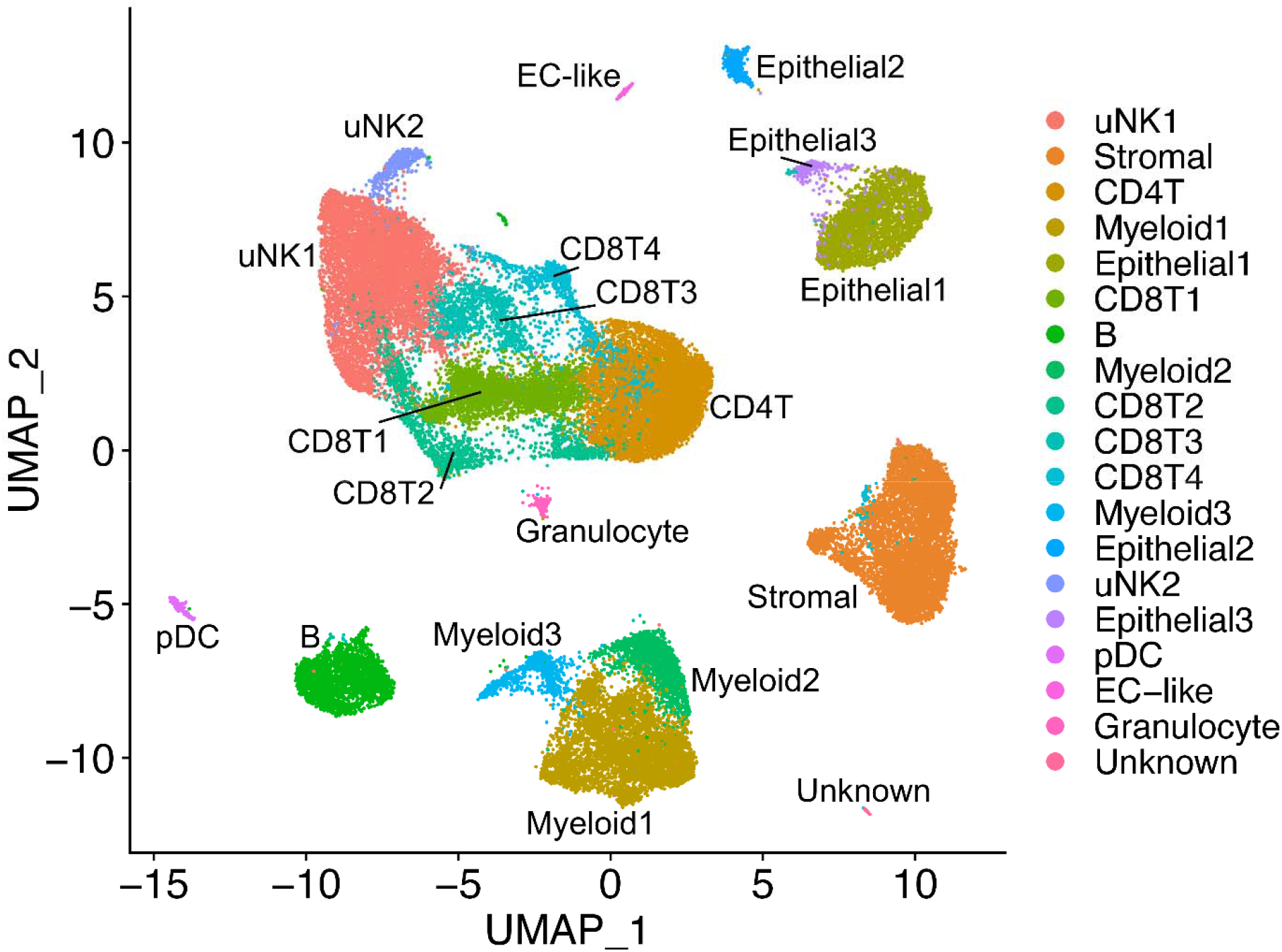
*Cellular composition of digested ME based on scRNA-Seq*. UMAP plot for all 33 digested menstrual effluent (ME) samples (controls=9; endometriosis cases=11; symptomatic cases=13). Several well-delineated cell clusters include a large cluster of uterine NK cells (uNK1), as well as clearly separated stromal cells, epithelial cells, and B cells. Several clusters each of T cells and myeloid cells are also defined, as well as a small cluster of plasmacytoid dendritic cells (pDC). A small cluster of approximately 60 unknown cells is in the lower right corner. The positive gene markers used to generate the cell clusters shown are included in Additional File 2: Table S1.

Seurat distinguishes multiple cell clusters shown on the UMAP (uniform manifold approximation and projection) plot. There is striking diversity of the cell types defined by the cluster analysis. A major group of uterine NK cells is designated cluster uNK1, with a small associated cluster designated uNK2. Sets of clusters related to CD8+and CD4+ T cells are shown in the central portion of the plot. Endometrial stromal cells and epithelial cells are identified in major clusters in the right side of the UMAP plot. Subclusters of endometrial stromal cells are described below in detail. Based on (31), Epithelial1 appears to be a mix of lumenal and glandular epithelial cells, Epithelial2 is comprised of ciliated epithelial cells, and Epithelial3 is a separate set of CD326-expressing cells that do not overlap with Epithelial 1 or Epithelial2. Distinct clusters of B cells and myeloid cells can also be delineated, along with a small cluster of plasmacytoid dendritic cells (pDC). The positive gene markers used to generate the cell clusters shown in Fig. 2 are included in Additional File 2: Table S1. Overall, the various cell clusters are well represented whether unfractionated whole ME or tissue-enriched ME is processed for scRNA-Seq. Some differences in cell subset frequencies can be observed; in particular, epithelial cells were enhanced when tissue-enriched ME was utilized for sample processing (see Additional File 3: Fig. S2).

### Cell clusters from ME containing endometrial tissue differ between endometriosis cases and healthy controls; relative depletion of uterine NK cells and enrichment of B cells in endometriosis cases

We compared the relative frequency of the various cell clusters in the freshly processed ME obtained from the diagnosed endometriosis cases (N=11) compared with controls (N=9), as shown in Fig. 3. By inspection of Fig. 3, it is apparent that both clusters of uNK cells (uNK1 and uNK2) are markedly depleted in the cases vs. controls (average percentage of uNK approximately 8% in cases, 28% in controls), as well as an increase in the proportion of B cells in cases (∼9%) vs. controls (∼3%). The odds ratios and confidence intervals for these two cell enrichment patterns are shown in Fig. 4, along with the patterns of enrichment of all the other major cell clusters. While there is some variation among many of the different cell clusters, a formal analysis shows the most striking differences are observed for uNK cells, which are enriched in controls (and depleted in cases; uNK1, *P* <10E-16; uNK2, *P* <10E-16), along with a relative enrichment in the proportion of B cells in the cases diagnosed with endometriosis (and relatively depleted in controls; *P* <10E-16). Note that the stromal cell cluster is not significantly different between cases and controls (*P* > 0.05).

**Fig. 3.**
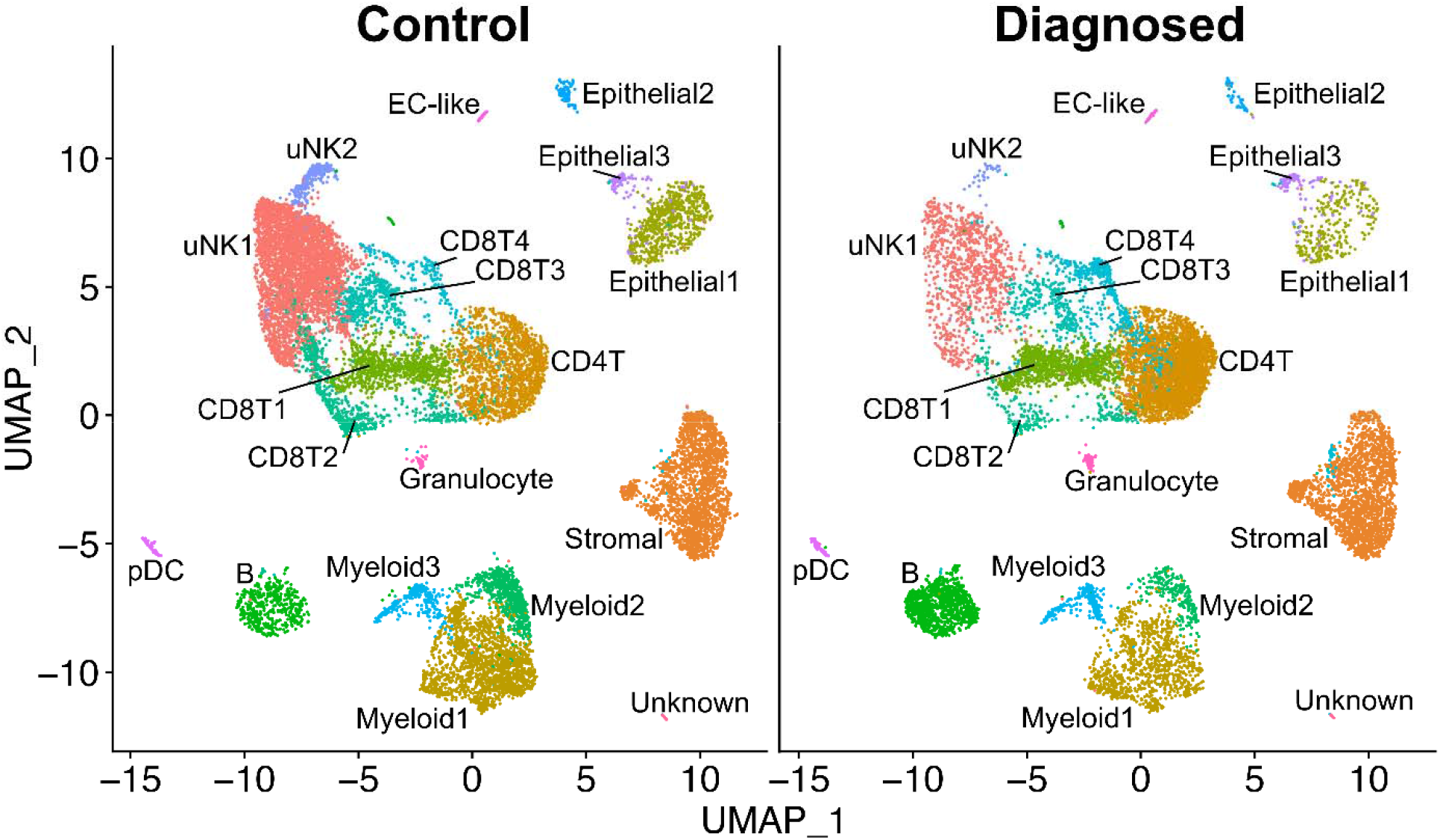
*Distinct cellular composition differences in digested ME from endometriosis cases vs. controls are revealed by scRNA-Seq*. The data taken from the UMAP plot in Fig. 2 is separated into two groups: controls (n=9, providing 14,327 cells) and endometriosis cases (n=11, providing 11,924 cells). The most striking difference is the increased fractions of uterine NK cells (uNK1 and uNK2) in the endometrial tissues of controls as compared to cases. In contrast, B cells are significantly enriched in cases. A formal analysis of enrichment is given in Fig. 4 and confirms the significant enrichment of uNK cells and B cells in controls and cases, respectively. The positive gene markers used to generate the cell clusters shown are included in Additional File 2: Table S1.

**Fig. 4.**
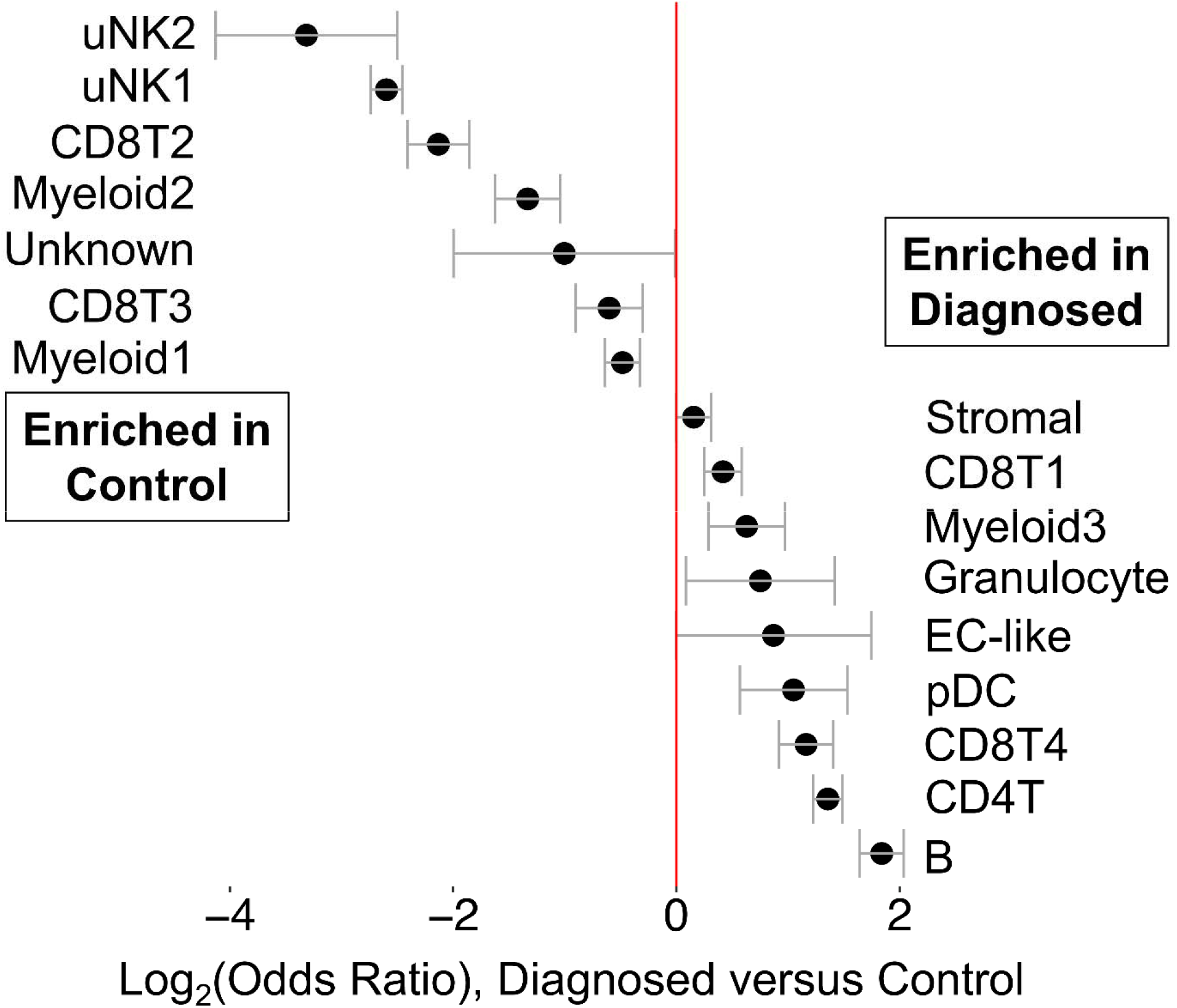
*Analysis of enrichment of cell subsets in ME comparing endometriosis cases and controls*. These data are taken from data shown in Fig. 3. The Log_2_ odd ratios (OR) with cell subsets enriched in controls on the left and cell subsets enriched in cases on the right. It is apparent that uterine NK (uNK) cells, both uNK1 and uNK2, are significantly enriched in controls, while B cells show the greatest enrichment in cases. Note: Epithelial cells are excluded from this analysis because their enrichment was affected by the tissue preparation method used.

We also explored whether the various proportions of cell clusters of the ME preparations from the “symptomatic” but undiagnosed group of subjects (N=13) are different from ME preparations from controls. This is clearly the case, as shown in Additional File 4: Fig. S3. Here, we show the relative enrichment of uNK cells is maintained in controls in comparison to the symptomatic group (uNK1, *P* <10E-16; uNK2, *P* = 0.0025), similar to that observed with ME from cases. B cells also show a significant relative enrichment in symptomatic as well as diagnosed cases, compared with controls (Additional File 4: Fig. S3) (symptomatic vs. control, *P* = 5.8 ×10^-6^), similar to that observed with ME from cases. Perhaps not surprisingly, these significant differences in symptomatic cases vs. controls are less striking than the differences in endometriosis cases vs. controls, given the likely heterogeneity of the symptomatic group.

### Decidualized stromal cell subclusters are reduced in endometriosis

Previous studies have reported reduced decidualization capacity in endometrial stromal cells grown from biopsies of patients with endometriosis (32). We have also observed impaired decidualization using stromal cells grown directly from ME (11, 15). Therefore, we examined whether this trend could be observed in fresh stromal cells analyzed by scRNA-Seq. The stromal cell numbers or percentages did not significantly differ between the control and endometriosis groups, as shown in Fig. 3 and Fig. 4. However, subclustering of the stromal cell cluster clearly identified 5 subclusters of interest within the stromal cell population (Fig. 5A). We have designated these subclusters based on the dominant transcripts expressed in each of these subclusters, as shown in the violin plots in Fig. 5B. Two of the five subclusters (2 and 4) are not different between cases and controls (the top genes of subclusters 2 and 4 are described in Additional File 5: Table S2). The subclusters showing significant enrichment in either cases or controls (subclusters 1, 3, and 5) are indicated by the Log_2_ (odds ratios, [OR]) below the UMAP plot (Fig. 5C).

**Fig. 5.**
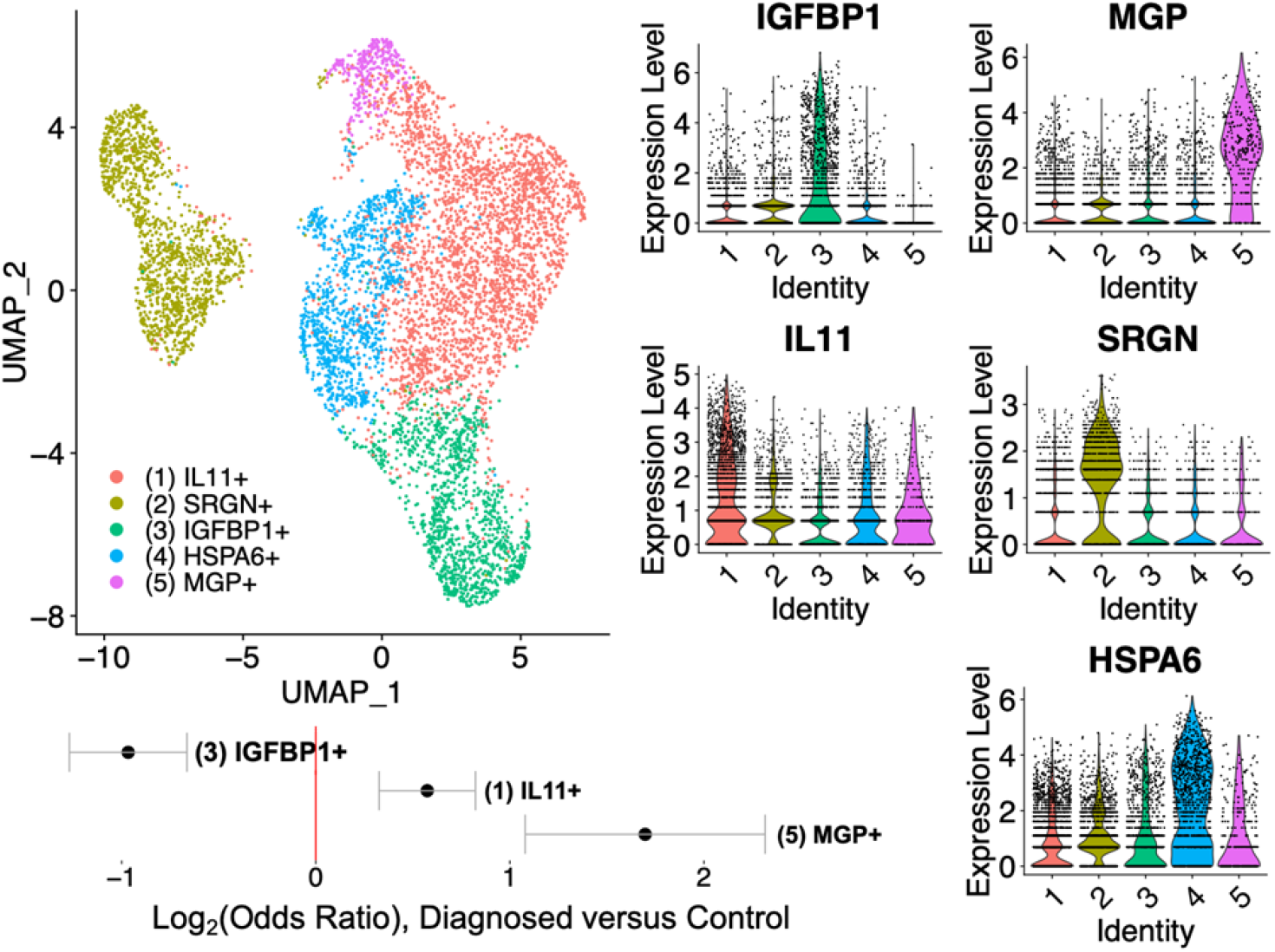
*Analysis of the stromal cell subclusters*. A) UMAP plot of the five stromal cell subclusters are shown. B) Violin plots showing the defining gene expression per subcluster for subclusters 1-5. C) Log_2_ (Odds Ratio) shows that subcluster 3 (IGFBP1+) is significantly enriched in controls (Log_2_ OR = -1.3, case vs. control). In contrast, subcluster 1 (IL11+) and subcluster 5 (MGP+) are enriched in diagnosed subjects. The top transcripts characterizing these three distinct stromal cell subclusters are summarized in Fig. 6 and emphasize the enrichment of the decidualized stromal cells – subcluster 3 (IGFBP1+) – in controls.

It is striking that an apparently decidualized stromal cell subcluster (expressing *IGFBP1* mRNA) is significantly enriched in controls compared with endometriosis cases (Fig. 5A-B). In addition to *IGFBP1*, the top differentially expressed genes in this subcluster (compared to other stromal cell subclusters) include *LEFTY2, DCN, LUM, MDK, C1QTNF6, APOE/D, DCN*, and other progesterone sensitive and decidualization/fertility gene markers (see left panel (subcluster 3) in Fig. 6 and Additional File 6: Table S3). (33-41) (42-46, 47, 48-103) (104-108) (109-113). This suggests that a phenotype of “decidualization” can be measured directly in stromal cells derived from fresh ME and is associated with control vs. disease phenotype. A modest enrichment of a subcluster expressing *IL11* was observed in cases, as indicated in Fig. 5A-C. In addition to *IL11*, this subcluster is associated with transcripts for *MMP3, MMP1, MMP9, SERPINB2, S100A6*, and *CXCL8*, among other genes associated with inflammation, fibrosis and senescence, as well as endometriosis, as shown in the middle panel (subcluster 1) of Fig. 6 (and Additional File 6: Table S3). A third subcluster, designated by high expression of the gene encoding matrix Gla protein (*MGP*), is also enriched in the stromal cells of cases (Fig. 5A-C). This subset expresses numerous extracellular matrix genes that have been associated with presence of perivascular stromal cells, senescence, and cell adhesion/cell spreading, including *FN1* (which encodes fibronectin-1), a known risk locus for endometriosis (114). Fig. 6 (right panel (subcluster 5) and Additional File 6: Table S3) also shows the list of top genes expressed in this subset. Additional File 7: Fig. S4 demonstrates that the *IGFBP1+* and *MGP+* subclusters map to stromal cells subsets defined in the decidua found in the first trimester of pregnancy by Vento-Tormo et al (26).

**Fig. 6.**
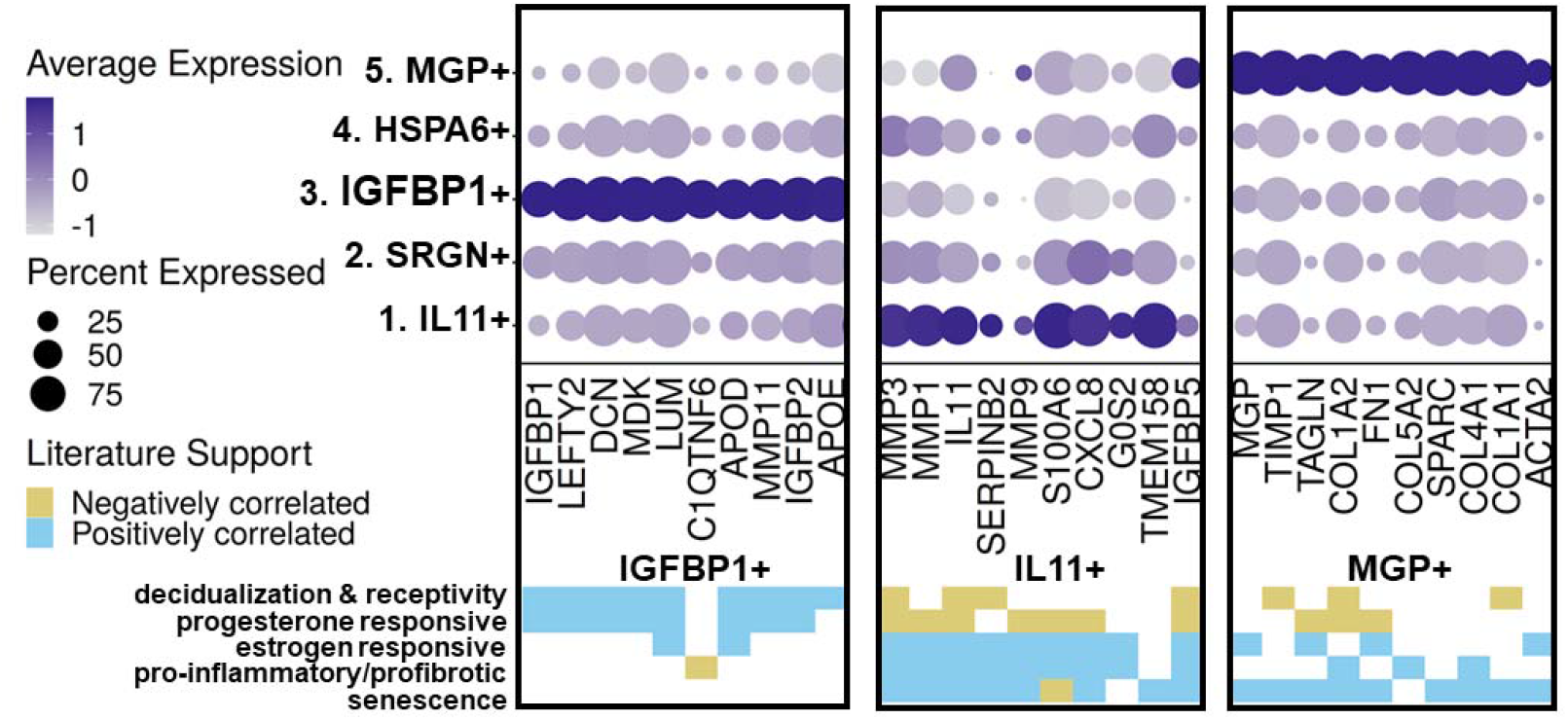
*Distinct subclusters of decidualized stromal cells and pro-inflammatory stromal cells distinguish ME from controls and endometriosis cases*. Upper panel: A summary of genes enriched in the stromal cell subclusters which are significantly enriched in cases (subclusters 1 [IL11+] and 5 [MGP+]) or controls (subcluster 3 [IGFBP1+]). Note: Subclusters 2 and 4 were not significantly different in cases vs. controls; see Additional File 6; Table S3 for the listing of genes differentially expressed in these clusters. Lower panel: Characteristic features of stromal cell subcluster gene markers. The decidualized stromal cell subcluster (IGFBP1+, subcluster 3) is prominently enriched in genes that are associated with decidualization and uterine receptivity and are progesterone responsive. In contrast, the non-decidualized stromal cell subsets that are enriched in cases (MGP+ [subcluster 5] and IL11+ [subcluster 1]) are variably enriched in estrogen responsive genes, and remarkably enriched in genes associated with inflammation, fibrosis, and cellular senescence. Note: MGP+ (subcluster 5) is also enriched in cell adhesion and cell spreading gene markers.

Finally, we examined the differences between cases and controls in the two uNK subclusters present in digested endometrial tissues in ME (uNK1 and uNK2, see Fig. 2). We noted a distinct subcluster of uNK cells (uNK2) that is characterized by the expression of genes associated with cell proliferation such as *MKI67* (which encodes Ki67) and *TOP2A* (which encodes topoisomerase 2A) (see Additional File 8: Fig. S5 for a full uNK subcluster analysis). As discussed below, this cluster also mapped nearly exactly (97%) with a proliferative subset of uNK cells that has been defined by scRNA-Seq in decidua obtained during the first trimester of pregnancy (26). This is consistent with the proliferation of uNK cells and overall accumulation of uNK cells in the course of decidualization in control subjects vs. cases, as shown in Fig. 3 and Fig. 4.

## Discussion

These studies show for the first time that the phenotype of eutopic endometrial tissue shed into the menstrual effluent is distinct in patients with endometriosis compared to control subjects. There are three major observations. First, the endometrial stromal cells show a relative deficiency of progesterone-sensitive gene markers associated with endometrial stromal cell decidualization in patients with endometriosis (e.g., *IGFBP1, LEFTY2, LUM, DCN,* etc). This is consistent with previous studies showing impaired decidualization of cultured endometrial stromal cells obtained from endometrial and ectopic endometriosis biopsies (32, 115), as well as from menstrual effluent (11, 15). Secondly, there is a striking reduction in the proportion of uNK cells in the ME-derived endometrial tissue of patients with endometriosis compared with controls. This was suggested by our previous studies of free cells present in ME using flow cytometry methods [11], but it is clearly a major distinguishing feature of the eutopic endometrium of endometriosis patients. Thirdly, our data suggest an enrichment of B cells in the eutopic endometrium of patients with endometriosis, a finding that is consistent with the hypothesis that chronic inflammation and/or chronic endometritis is a predisposing factor in the development of endometriosis (116).

A deficiency in the decidualization capacity of stromal cells cultured from biopsies of the eutopic endometrium has been reported previously (32), and is also found in ME-derived stromal cells collected at the time of menstruation (11, 15). Our scRNA-Seq data clearly shows the reduction of the *IGFBP1+*-expressing decidualized stromal cell subclusters in endometriosis cases vs. controls (Fig. 5C). The relationship of this finding to the pathogenesis of endometriosis is not established. One possibility is that this differentiation deficiency leaves behind non-decidualized endometrial stromal cells that exhibit proinflammatory, pro-fibrotic, and/or senescent phenotypes. These ‘pathogenic’ cells may then initiate or promote lesions following retrograde transfer into the peritoneal cavity. The enrichment of an *IL11*-expressing stromal cell subcluster in the endometriosis ME samples that express many estrogen-responsive, pro-inflammatory, pro-fibrotic and senescence gene markers (shown in Fig. 6 and Additional File 6: Table S3) provides some support for this possibility, but this needs confirmation in larger datasets. The significant increase in the *MGP*+ stromal subcluster in endometriosis (Fig. 5C) is also of potential interest. As shown in Fig. 6 (right panel), the *MGP*+ stromal cell subcluster expresses many genes that are associated with the extracellular matrix, including *FN1* (encoding fibronectin-1) which has been associated with an increased risk for endometriosis in GWAS studies (117). Interestingly, most of the top markers found in the *IL11+* and the *MGP*+ subclusters are either associated with senescence or induce senescence (e.g., *IL11* and *SERPINB2* [Fig. 6 and Additional File 6: Table S3]). Inflammation and senescence are key features of endometriosis and reduced uterine receptivity and infertility (118-120).

Another possibility is that the overall environment of the eutopic endometrium predisposes to reduced stromal cell decidualization, independent of any direct role or effect on stromal cell subsets in the disease. A chronic inflammatory endometrial environment might lead to, or be associated with, other changes that put individuals at risk for endometriosis. For example, the presence of chronic endometritis has been reported to be a significant risk factor for endometriosis (116, 121); chronic endometritis is also associated with reduced stromal cell decidualization (122). Interestingly, the presence of B cells in endometrial tissue, particularly plasma cells, is a requirement for the clinical diagnosis of chronic endometritis (116). We note the significant increase in B cells in shed endometrium of endometriosis patients (Fig. 3 and 4) and symptomatic subjects (Additional File 4: Fig. S3) when compared to controls. This may reflect an inflammatory state, as B cells play an important role in mediating or regulating inflammatory and autoimmune diseases (123). The numbers of B cells available for detailed analysis have not allowed us to fully understand the phenotype of these cells; this is an area for future study.

We have demonstrated that uNK cells are remarkably depleted in the ME-derived endometrial tissues of patients with endometriosis (Fig. 3 and Fig. 4 and Additional File: Fig. S3). This may reflect compromised decidualization in these subjects. uNK cells are a characteristic feature of decidualizing tissues (124) and are also prominent in the decidua of early pregnancy (26). To our knowledge, this is the first report of proliferating uNK cells found in ME. Crosstalk between stromal cells and uNK cells is a feature that promotes decidualization and uterine receptivity/placental vascular remodeling (125). uNK cells do not appear to play a major role in decidualization in uNK deficient IL15 knockout mice (126). It remains unclear whether uNK cells or stromal cells are the primary driver of the decidualization impairment in endometriosis.

However, uNK cells do play a role in the maintenance of decidual integrity as reported by Ashkar et al (127). Brighton and co-workers emphasized the important role of uNK cells in clearing senescent decidual cells in the cycling human endometrium and their clearance is proposed to be important for optimal fertility (128). A lack of uNK cells in the endometrium may contribute to increased numbers of senescent cells observed in the stromal subclusters among endometriosis subjects and may contribute to endometriosis-associated infertility. However, it is plausible that a lack of decidualizing endometrial stromal cells (with concomitant reduced production of IL-15 and uNK chemoattractants) reduces the infiltration and proliferation of uNK in decidualizing zones. Defective uNK cell function has recently been proposed in the setting of endometriosis with infertility (129). We did not observe *IL15* expression by ME-stromal cells; this is not surprising as IL-15 expression by stromal cells peaks before the mid-secretory phase.

Interestingly, we did observe enhanced expression of *IL2RB* (which encodes a component of the IL-15 receptor) in the uNK cells of controls compared with the endometriosis group. Since uNK cells are reported to play a role in infertility (124, 130), and infertility is a common feature of endometriosis, further analysis of the uNK subset will clearly be of interest.

Taken together, these observations suggest a set of interactions that may drive the development and/or progression of endometriosis at several levels, as summarized in Fig. 7. Stromal cell decidualization may be inhibited by a number of factors, including chronic inflammation, stress and/or progesterone resistance. This may divert stromal cells into a more proinflammatory/senescent state. Furthermore, deficient decidualization may also compromise the infiltration of uNK cells into the decidua, and therefore reduce the clearance of senescent cells. Clearly, host genetic variation may influence these processes at every level of these interactions.

**Fig. 7.**
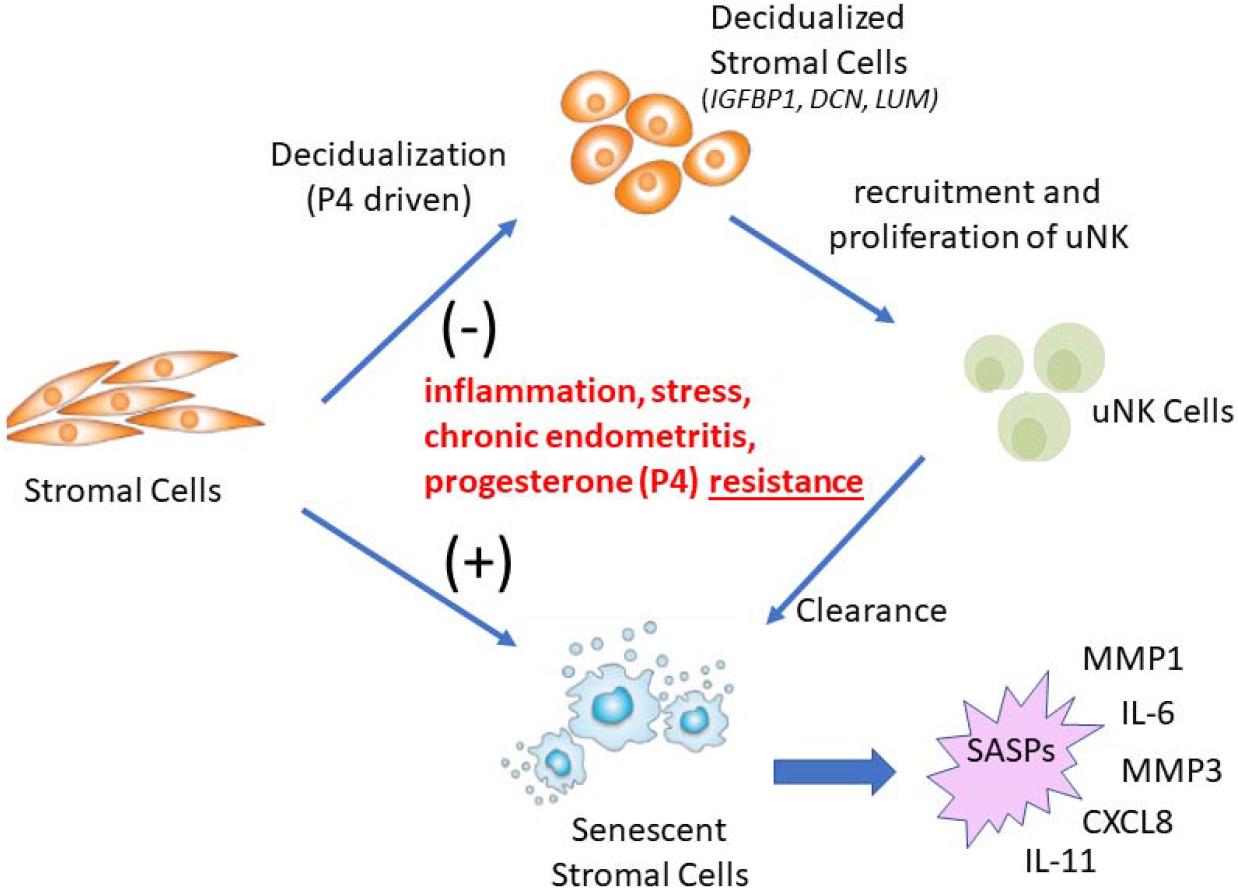
*A disease model for endometriosis*. Defective endometrial stromal cell decidualization may be driven by multiple factors including inflammation, chronic endometritis, stress, and/or progesterone resistance. This, in turn, may direct stromal cell differentiation in the direction of chronic inflammation and senescence, with accompanying senescence associated secretory phenotypes (SASPs), which include pro-inflammatory mediators and proteases. The senescence phenotype may also impair decidualization. Reduced decidualization may also compromise the infiltration and proliferation of uNK cells, which are likely to be important for senescent cell removal. Further analysis of other cells in menstrual effluent will be important to provide further support for this model.

It is encouraging that many of our findings in patients with pathologically confirmed endometriosis are also present in a proportion of subjects with chronic symptoms that are suggestive of endometriosis, even in the absence of a confirmed tissue diagnosis. The delay in diagnosis of endometriosis is widely recognized as a major barrier in the management of this disease, with delays of up to a decade in some subjects before the disease is recognized (131). We recognize the limitation that the symptomatic group lacks a diagnosis and therefore, we cannot assess the predictive ability of our results. To address this limitation, a clinical trial is underway to enroll symptomatic subjects who are being evaluated by diagnostic laparoscopy as part of their standard care by collaborating surgeons; scRNA-Seq profiles of their ME collected prior to surgery will be validated based on the results of their laparoscopic diagnosis. Such a study design will be required to establish the positive and negative predictive value of menstrual tissue analysis in a real-world clinical setting where an endometriosis screening test might be applied.

Due to the cost and complexity of the analysis, an scRNA-Seq approach is unlikely to become a diagnostic test for endometriosis. However, we propose that the data obtained from scRNA-Seq can be leveraged to develop future diagnostic and/or screening tests. What should such screening tests involve? It will likely include an assessment of gene expression patterns among ME-derived stromal cells or uNK cells (or specific stromal cell and uNK subsets). An initial analysis of stromal cell clusters suggests several potentially useful gene expression differences among cases vs. controls (Additional File 9: Fig. S6). Differences are also observed in uNK cells (Additional File 10: Fig. S7) or indeed may be found in other cell types as well.. On the other hand, if it can be adapted to a clinical diagnostic test, scRNA-Seq of these tissues is likely to be the most informative approach, perhaps having more global utility to establish complex and heterogenous disease subtypes, as well as predicting or following response to therapy.

Additional phenotypes that can be uncovered using scRNA-Seq analysis on larger populations may yet yield additional biomarkers that can be incorporated into a more targeted multivariate biomarker analysis for diagnostic purposes.

In any case, the integration of our findings into a unified picture of the pathogenesis of endometriosis will require additional scRNA-Seq studies of larger heterogenous populations, at different stages of disease development and include deeper analysis of T cells, B cells, myeloid cells, and epithelial cells. Abnormalities of the eutopic endometrium are widely recognized features of endometriosis [6-9, 18, 34] and this can provide diagnostic value, regardless of whether retrograde menstruation plays a causative role. In addition, it is likely that scRNA-Seq approaches of the endometrium via ME may allow for improved classification of clinically meaningful disease subsets and as a means for assessing patients’ responses to therapies, as well as uterine-associated fertility status. For example, many of the genes that exhibit changes in the stromal cell subclusters are associated with either estrogen or progesterone responsiveness (Fig. 6), and these differences could be used to guide or assess responses to hormonal therapies and for assessing aspects of uterine receptivity/fertility.

On the other hand, if disease causation is due to retrograde menstruation of abnormal endometrial tissues, ME analysis provides an opportunity to explore new therapies. For example, based on the enrichment of pro-senescent genes in endometriosis endometrial stomal cells (vs. control cells) and the deficit of uNK cells in endometriosis subjects, we propose investigating senescence as a central feature of endometriosis. Once demonstrated, this may have important potential therapeutic implications, since various senotherapeutics (senolytic and senomorphic agents) have now been shown to improve chronic inflammatory diseases in pre-clinical models and human clinical trials (132, 133). This is significant since none of the current medical therapies for endometriosis have been shown to alter disease progression.

## Conclusions

In summary, these scRNA-Seq data of ME collect from endometriosis cases and healthy controls represent a first attempt to globally characterize the cellular diversity of endometrium that is shed at the time of menstruation. More detailed studies in larger datasets are clearly required, particularly regarding diversity in T cells, B cells and myeloid cells, as well as epithelial cells. We propose that a comprehensive assessment of cellular phenotypes in ME tissues will open a new window on both diagnosis as well as preventive treatment for patients at risk for endometriosis as well as other uterine and reproductive disorders.

## Data Availability

All data produced in the present study are available upon reasonable request to the authors

## Abbreviations

BSA: Bovine serum albumin
DTT: Dithiothreitol
FBS: Fetal bovine serum
GEO: Gene Expression Omnibus
H&E: Hematoxylin and eosin
MASC: Mixed-effects modeling of associations of single cells
ME: Menstrual effluent
nUMI: Number of unique molecular identifiers
OR: Odds ratios
pDC: Plasmacytoid dendritic cells
SASP: Senescence associated secretory phenotype
scRNA-Seq: Single cell RNA-sequencing
SD: Standard deviation
SSC: Saline sodium citrate
UMAP: Uniform manifold approximation and projection
uNK: Uterine natural killer

## Declarations Acknowledgements

We are grateful to the Endometriosis Foundation of America and to Dr. Tamer Seckin for providing early support for this work, and the for the ongoing support from the Northwell Health Innovation Award. We also thank Anthony Liew, Cassie Pond and Maruf Chowdhury who provided valuable technical support for this project. We are especially grateful to the many extraordinary patients and volunteers without whose participation this project could not have been accomplished.

## Ethics Approval and Consent to Participate

All procedures for the collection of samples from research subjects were performed with the approval of the institutional review board (IRB) of the Feinstein Institutes/Northwell Health. All participants signed informed consent prior to enrollment and study participation.

## Consent for Publication

All authors give their consent for publication of this manuscript.

## Availability of Data and Materials

The original data and materials presented in the study are available from the corresponding authors upon reasonable request. scRNA-Seq data is available at National Center for Biotechnology Information/Gene Expression Omnibus (GEO) (accession number GSE203191).

## Competing Interests

The authors declare that they have no competing interests.

## Funding

This work was supported by the Northwell Health Innovations Award and the Endometriosis Foundation of America

## Authors’ Contributions

Conceptualization: CNM, PKG. Recruitment and enrollment: KE, MDF. Data collection: AJS, RPA, KE, HK, MDF. Formal analysis: PKG, CNM, AJS, RPA. Sample processing: RP, PKC, HV, RH, AN. Pathology slide review: AMT. Library construction: HK. Funding acquisition: CNM, PKG. Supervision of scRNA-Seq: ATL. Writing—original draft: PKG, CNM, AJS, RPA. Writing— review & editing: HK, PKG, CNM, RPA, AJS. All authors read and approved the final manuscript.

## Notes

### Competing Interest Statement

The authors have declared no competing interest.

### Author Declarations

Ethics Committee/IRB of Feinstein Institutes at Northwell Health gave ethical approval for this work

### Summary of Updates

this version of the manuscript includes a model for endometriosis pathogenesis shown in new Figure 7

